# The impact of prior COVID-19 on vaccine response and the resultant hybrid immunity are age-dependent

**DOI:** 10.1101/2022.09.19.22280079

**Authors:** Sachie Nakagama, Yu Nakagama, Yuko Komase, Masaharu Kudo, Takumi Imai, Yuko Nitahara, Natsuko Kaku, Evariste Tshibangu-Kabamba, Yasutoshi Kido

## Abstract

**Background:** More people with a history of prior infection are receiving SARS-CoV-2 vaccines. Understanding the magnitude of protectivity granted by ‘hybrid immunity’, the combined response of infection- and vaccine-induced immunity, may impact vaccination strategies.

**Methods:** A total of 36 synchronously infected (‘prior infection’) and, 33 SARS-CoV-2 naïve (‘naïve’) individuals participated. Participants provided sera six months after completing a round of BNT162b2 vaccination, to be processed for anti-spike antibody measurements and neutralization assays. The relationships between antibody titer, groups and age were explored.

**Results:** Anti-spike antibody titers at 6 months post-vaccination were significantly higher, reaching 13- to 17-fold, in the ‘prior infection’ group. Linear regression models showed that the enhancement in antibody titer attributable to positive infection history increased from 8.9- to 9.4- fold at age 30 to 19- to 32-fold at age 60. Sera from the ‘prior infection’ group showed higher neutralizing capacity against all six analyzed strains, including the Omicron variant.

**Conclusions:** Prior COVID-19 led to establishing enhanced humoral immunity at 6 months after vaccination. Antibody fold-difference attributed to positive COVID-19 history increased with age, possibly because older individuals are prone to symptomatic infection accompanied by potentiated immune responses. Durable protection of hybrid immunity deserves reflection in vaccination campaigns.

## Introduction

As the cumulative incidence of COVID-19 increases worldwide, more people with a history of prior infection are now receiving SARS-CoV-2 vaccines. With the infection-induced and vaccine-induced immune responses having different protective characteristics,^1^ the acquisition of such a combined immune response is drawing attention as ‘hybrid immunity’. Understanding the magnitude of protectivity against SARS-CoV-2 granted by hybrid immunity and its role in the establishment of herd immunity may impact future vaccination strategies.

With immunopotentiation through repeat vaccinations becoming a pivotal strategy, a consensus ought to be reached on the target population, optimal interval, and dosing regimen for the repeated boosters. To accomplish this, it is becoming increasingly important to understand the longitudinal evolution of the antibody response and the resulting ‘residual immunity’ following vaccination dose(s). The impact of prior infection on the acquisition of protective immunity in vaccinated individuals has been actively studied since the introduction of the SARS-CoV-2 vaccines.^2^ However, possibly due partly to adherence challenges, many studies have focused on the differences in the early-phase post-vaccine response between naïve and previously infected individuals,^3,4^ whereas fewer studies have described this in the mid- to long-term.

We previously carried out a SARS-CoV-2 seroprevalence survey targeting healthcare workers (HCWs) from a tertiary care hospital in Japan. This revealed a nosocomial cluster infection of which the burden had been underestimated, accumulating up to 15.5% overall seroprevalence.^5,6^ Through longitudinal follow-up and further serological description of the cohort of HCWs,^7^ we took advantage of the opportunity to investigate a uniformly conditioned population endowed with hybrid immunity: those synchronously infected through a nosocomial cluster infection, and again synchronously administered the BNT162b2 vaccine through the nation’s mass vaccination campaign. The incremental effect of hybrid immunity on an individual’s long-term residual antibody titers was analyzed. These observations suggest the need to rethink our vaccination campaign strategies that currently recruit and treat those with prior infections and those without equally.

## Materials and Methods

### Participants and serum sampling

The participants in this study were HCWs at the St.Mariannna University, Yokohama Seibu Hospital, Kanagawa, Japan, where we previously conducted an anti-SARS-CoV-2 seroprevalence survey.^5^ In the previous study, 64 COVID-19-affected HCWs and 350 non-infected individuals were identified following an outbreak having occurred in the hospital during April–May 2020. From the cohort, 36 individuals who had tested positive (‘prior infection’) and 33 individuals who had tested negative (‘naïve’) on Roche Elecsys anti-SARS-CoV-2 (Roche Diagnostics, Rotkreuz, Switzerland) antibody testing agreed to participate in this follow-up study. The ‘naïve’ individuals were further confirmed to have negative anti-nucleocapsid serology upon study entry. Those categorized as the ‘prior infection’ group, as HCWs, were kept under continuous health monitoring and were confirmed to have had no signs or symptoms indicative of COVID-19 re-infection since completion of the previous survey until their enrollment in this present study.

All participants received two doses of the BNT162b2 vaccine at the standard three-week interval, according to the recommended vaccination schedule in Japan. Participants provided their sera six months after completion of their second BNT162b2 dose, during the period of November 15-24, 2021 (with only two exceptions, each providing their sera four and five months after completion). The donated sera were processed for anti-spike antibody titer measurements and neutralization assays.

The study was approved by the Osaka Metropolitan University Institutional Ethics Committee [#2020-003]. Written consent for participation was obtained from every participant.

### Assessment of anti-spike humoral immunity

The anti-spike antibody titer was measured using two fully automated, commercially available immunoassay platforms. The chemiluminescence immunoassay, Abbott SARS-CoV-2IgG II Quant (Abbott Laboratories, IL, USA), was designed to detect serum IgG antibodies targeting the spike protein of SARS-CoV-2. The electrochemiluminescence (ECL) immunoassay, Roche Elecsys anti-SARS-CoV-2 S (Roche Diagnostics, Rotkreuz, Switzerland), was designed to detect serum total antibodies targeting the spike protein. The assays were performed according to the manufacturers’ instructions.

### Evaluation of the neutralizing capacity of anti-SARS-CoV-2 antibodies

1:10 diluted serum samples were tested with the Meso Scale Discovery neutralization assay, an ECL-labeled competition immunoassay. The V-PLEX SARS-CoV-2 Panel 22 (ACE2) Kit (K15562U) (Meso Scale Diagnostics LLC, MD, USA), containing spots coated with Wuhan, Alpha, Beta, Delta, Gamma, and Omicron antigens, evaluated the capacity of serum anti-SARS-CoV-2 antibodies to inhibit the receptor binding domain-ACE2 binding. The ECL signal, negatively proportional to the concentration of neutralizing antibodies in the sample, was read on the MESO QuickPlex SQ 120MM instrument (Meso Scale Diagnostics LLC). Neutralizing capacity was calculated from the following formula and was expressed as ‘Inhibition rate (%Inhibition)’: %Inhibition = { 1 – (ECL signal of sample) / (ECL signal of blank)} × 100 [%].

### Statistical Analysis

Participants’ demographics were described as numbers (and/or percentages) for categorical variables and as means ± standard deviation for continuous variables, and were compared between ‘naïve’ and ‘prior infection’ groups by the chi-square test or the Mann-Whitney’s U test. The antibody titer was expressed as geometric mean titer (GMT) [95% confidence interval] and compared between groups by the t-test on a logarithmic scale. The relationships between antibody titer, groups (‘naïve’ and ‘prior infection’) and age were explored using linear regression models. The age-specific ratios of GMT were estimated based on t-distribution. The dimorphism of age effect on the log-transformed post-vaccination antibody titer was examined by ANCOVA, testing for interaction between groups and age. The distributions of %Inhibition in ‘naïve’ and ‘prior infection’ groups were expressed as medians [interquartile ranges] and compared by the Mann-Whitney’s U test. P-values less than 0.05 were considered statistically significant.

## Results

A total of 69 participants (33 categorized as the ‘naïve’ group and 36 as the ‘prior infection’ group) were included in the analysis (Table 1). The cohort had a sex ratio of 87% female (88% in naïve vs. 86% in ‘prior infection’; P = 0.83) and a mean age of 42 ± 12 years (47 ± 9 years in ‘naïve’ vs. 37 ± 12 years in ‘prior infection’; P = 0.0005). Participants self-reported no pre-existing medical conditions known to critically affect antibody response towards any vaccine (i.e. diabetes mellitus, malignant disease, chronic kidney disease). Within the ‘prior infection’ group, the previous COVID-19 diagnosis was often a mild-to-moderate illness, except for a single case of severe disease. Anti-nucleocapsid antibodies remained negative in all ‘naïve’ throughout and remained above the positivity threshold in all of those with ‘prior infection’ except one who sero-reverted during the 20-month follow-up period.

**Table 1.** Descriptive characteristics of participants

| Variable | Naïve (n = 33) |  | Prior infection (n = 36) |  |
| --- | --- | --- | --- | --- |
| Age, y [SD]<sup>a</sup> | 47 | [9] | 37 | [12] |
| Sex, female [%] | 29 | [88] | 31 | [86] |
| Medical history, n [%] | 0 | [0] | 0 | [0] |
| Previous COVID-19, n [%] |  |  |  |  |
| Mild | - | [-] | 30 | [83] |
| Moderate | - | [-] | 5 | [14] |
| Severe | - | [-] | 1 | [3] |

<sup>a</sup> standard deviation

Compared with the ‘naïve’ group, anti-spike antibody titers at 6 months post-vaccination were significantly higher in the ‘prior infection’ group (Figure 1) (Abbott Architect anti-spike IgG titer 710 [537–939] vs. 9123 [6982–11921] AU/mL; P < 0.0001, Roche Elecsys anti-spike total antibody titer 480 [345–669] vs. 8168 [5945–11222] U/mL; P < 0.0001). For each immunoassay, there was an approximate 13- and 17-fold change, respectively, in the GMT ratio between groups.

**Figure 1.**
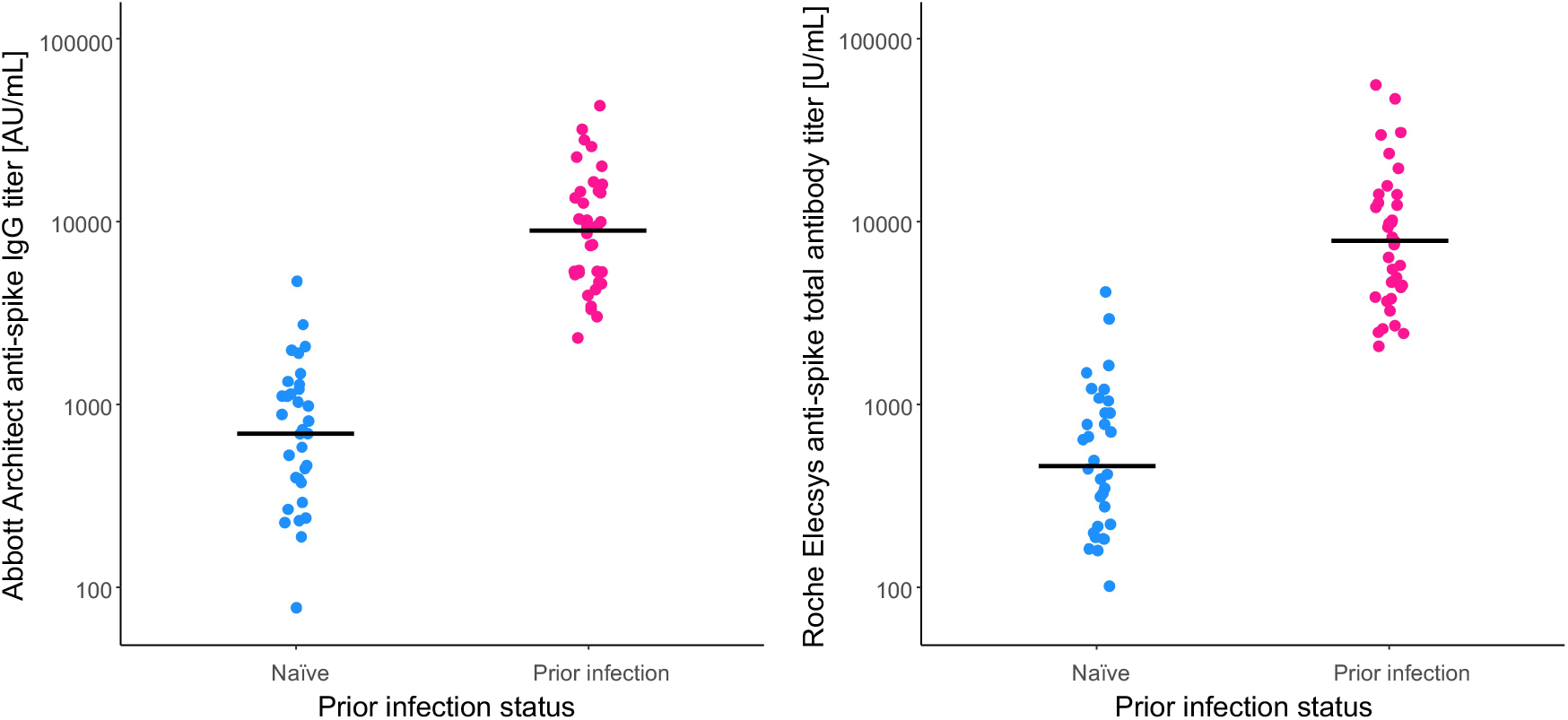
Anti-spike antibody titers after BNT162b2 vaccination. For comparison by prior infection status, the (A) Abbott Architect anti-spike IgG titers, and the (B) Roche Elecsys anti-spike total antibody titers at 6 months post-vaccination are shown with their respective geometric means (Solid lines).

Age was negatively associated with post-vaccination antibody titer in the ‘naïve’ group, whereas it was positively associated in the ‘prior infection’ group (Spearman’s correlation coefficients for Abbott and Roche titers, respectively: -0.20 and -0.25 in ‘naïve’, 0.38 and 0.52 in ‘prior infection’). Therefore, the impact of age on the differences in post-vaccination antibody titers was compared between the groups. Evaluated from linear regression models (Figure 2), the dimorphic effect of age on the log-transformed post-vaccination antibody titer was significant (P = 0.049 and 0.007, for Abbott and Roche titers, respectively). Interpolation from the regression models showed that the fold change in the GMT ratio increased from 8.9-fold at age 30 years to 19-fold at age 60 years for the Abbott IgG titer, and 9.4-fold at age 30 years to 32-fold at age 60 years for the Roche total antibody titer (Table 2).

**Figure 2.**
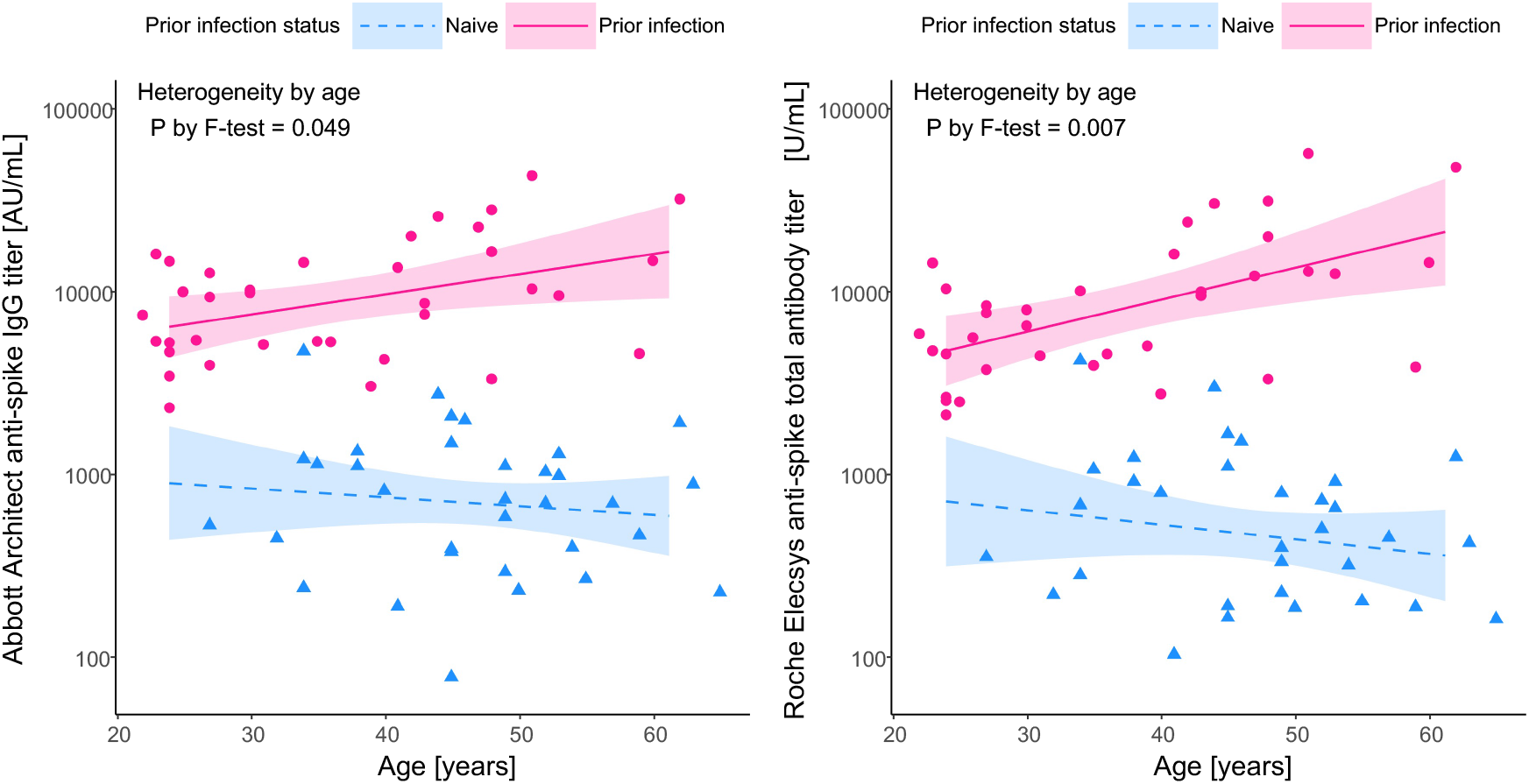
Age-dependent increase in the between-group (‘prior infection’ vs. ‘naïve’) differences in post-vaccination anti-spike antibody titer. The impact of age on the log-transformed anti-spike antibody titer is fitted with linear regression models. Solid and dashed lines represent the predicted anti-spike antibody titer calculated by the model for the (A) Abbott Architect anti-spike IgG assay and the (B) Roche Elecsys anti-spike total antibody assay. Shadowed areas represent the 95% confidence interval.

**Table 2.** Age-specific differences in Abbott Architect anti-spike IgG titers attributable to prior infection status

| Age, y | Ratio of GMT<sup>a</sup>, 'prior infection' to 'naïve' | [95% CI]<sup>b</sup> | P-value |
| --- | --- | --- | --- |
| Overall | 12.8 | [8.7–18.9] | < 0.001 |
| 30 | 8.9 | [4.7–16.8] | < 0.001 |
| 40 | 11.5 | [6.2–21.3] | < 0.001 |
| 50 | 14.8 | [7.6–29.2] | < 0.001 |
| 60 | 19.1 | [8.7–42.3] | < 0.001 |

<sup>a</sup> geometric mean of titer
<sup>b</sup> confidence interval

**Table 3.** Age-specific differences in Roche Elecsys anti-spike total antibody titer attributable to prior infection status

| Age, y | Ratio of GMT<sup>a</sup>, 'prior infection' to 'naïve' | [95% CI]<sup>b</sup> | P-value |
| --- | --- | --- | --- |
| Overall | 17.0 | [10.8–26.9] | < 0.001 |
| 30 | 9.4 | [4.6–19.5] | < 0.001 |
| 40 | 14.1 | [7.0–28.6] | < 0.001 |
| 50 | 21.1 | [9.7–45.7] | < 0.001 |
| 60 | 31.5 | [12.7–78.0] | < 0.001 |

<sup>a</sup> geometric mean of titer
<sup>b</sup> confidence interval

In the neutralization assay (Figure 3), sera of participants from the ‘prior infection’ group showed higher neutralizing capacity against all six strains, including the wild type (81.1 [61.1–91.5] vs. 99.8 [99.7–99.9] %; P < 0.0001), and the Alpha (68.1 [54.4–84.8] vs. 99.8 [99.6–99.8] %; P < 0.0001), Beta (38.4 [6.9–55.1] vs. 99.2 [97.0– 99.5] %; P < 0.0001), Gamma (51.1 [38.1–68.7] vs. 99.6 [98.4–99.8] %; P < 0.0001), Delta (78.2 [57.8–83.9] vs. 99.8 [99.7–99.9] %; P < 0.0001), and Omicron variants (0.0 [0.0–18.3] vs. 74.1 [39.4–84.9] %; P < 0.0001).

**Figure 3.**
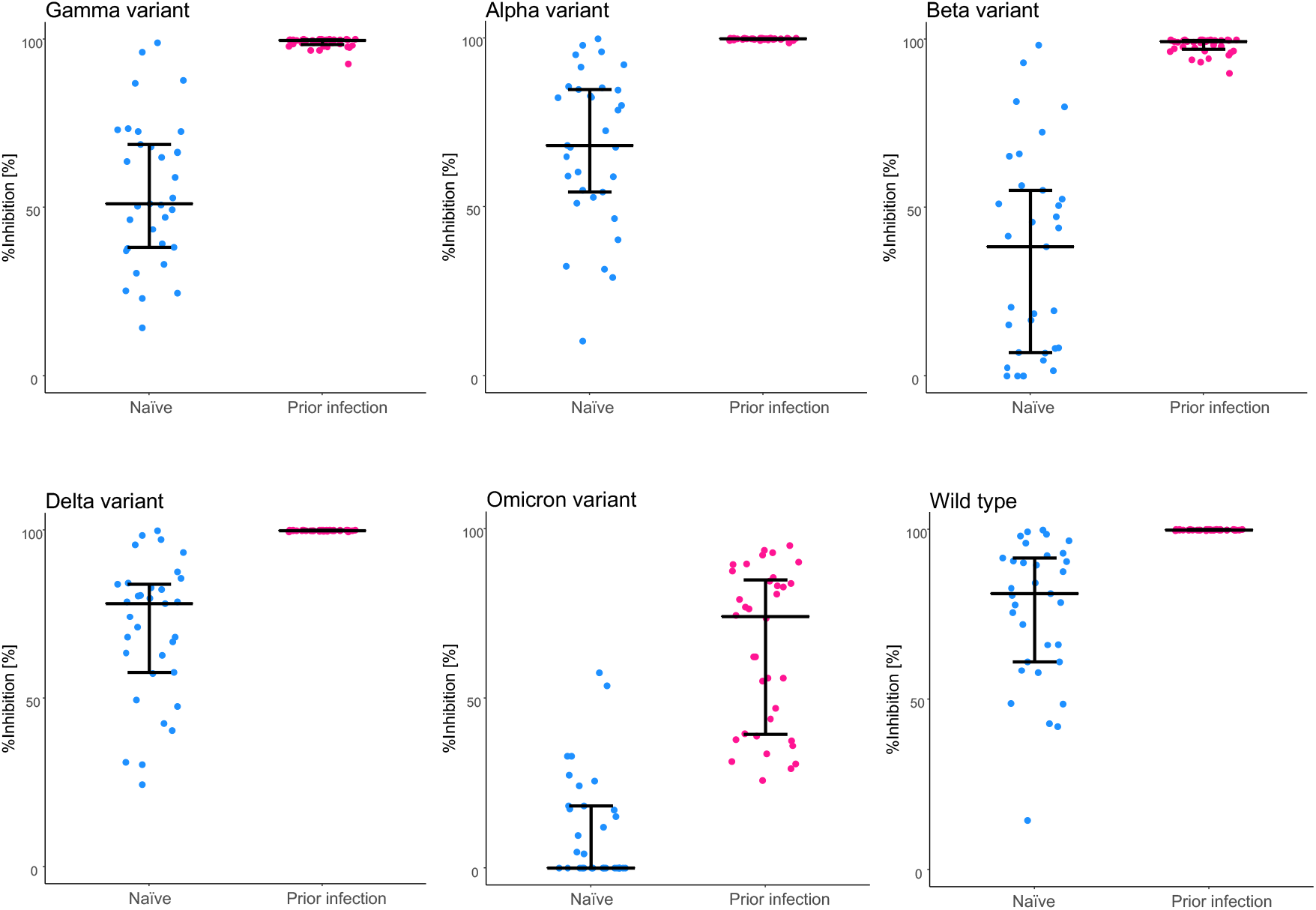
Neutralizing capacity of serum antibodies after BNT162b2 vaccination. For comparison of the variable level of protection granted depending on prior infection status, neutralizing capacity against the wild type and the variant SARS-CoV-2 spike antigen was assessed at 6 months post-vaccination. The bars (error bars) indicate medians (interquartile ranges). %Inhibition, inhibition rate.

## Discussions

The present study showed that prior infection was predictive of enhanced and durable residual immunity against SARS-CoV-2 at 6 months after vaccination. The magnitude of the difference in the antibody titer between the ‘prior infection’ and ‘naïve’ groups was age-specific and increased with older age. The superiority of ‘prior infection’ maximized at age 60 years, showing 19- and 32-fold higher Abbott and Roche antibody titers, respectively.

The IgG response following SARS-CoV-2 BNT162b2 vaccination (i) peaks rapidly within the first 2 months from the initial dose and then (ii) enters a subsequent stage of gradual decay.^8^ The initial studies reporting the effect of prior infection on BNT162b2 post-vaccination antibody titers had often targeted the peak response. At 2 months and 3 months after the initial dose, 3.7-fold and 2.7-fold increases, respectively, were observed in vaccinees with prior COVID-19 infection compared with the naïve group.^3,4^ While potentiation of the peak response to BNT162b2 vaccination by ‘prior infection’ has been well supported by abundant real-world data, the stage of IgG decay has been less addressed. Recently, a modeling study of post-vaccination ‘waning immunity’ showed that the anti-SARS-CoV-2 IgG levels of vaccinees with prior infection decreased at a slower rate compared to the non-previously infected.^9^ Another study also suggested a slower decay of antibody titers in the prior-infection group, resulting in a further exaggerated fold change in titer during the decay phase of antibodies.^10^ The here observed unexpectedly large 13- to 17-fold change in antibody titers, attributed to prior infection status, is thus fully interpretable considering the biphasic kinetics of the post-vaccination immune evolution.

Interestingly, age had dimorphic effects on post-vaccination immune evolution depending on prior infection status. Older age was associated with a higher level of IgG in previously infected individuals, whereas it was associated with a lower level of IgG in the naïve group. This can be explained by the fact that older individuals are more prone to symptomatic, and possibly more severe, SARS-CoV-2 infection, which in turn is often accompanied by a potentiated circulating IgG response.^5^ To support this idea, the present cohort of vaccinees with prior infection showed a strong positive correlation between the peak anti-spike antibody response following their COVID-19 diagnosis (at 2 months’ convalescence) and the residual antibody titer at 6 months post-vaccination (Pearson’s correlation coefficient: 0.71 and 0.77 for Abbott and Roche titers, respectively).

Immunopotentiation through repeated boosters is an affordable strategy only when the risk-benefit balance is optimized and deemed favorable. For the influenza vaccine, prior-year vaccination has shown to have negative effects on the current year’s vaccine effectiveness.^11^ Further, a frequent vaccination history was associated with 41% and 27% decreases in vaccine effectiveness against type A influenza and type B influenza, respectively.^12^ This phenomenon has been explained as ‘antibody feedback’.^13^ Potential ‘antibody feedback’ has also been suggested with the SARS-CoV-2 vaccines.^14^ An extended 3-month interval regimen has resulted in, on average, 3.5-fold higher IgG titers.^15^ A longer interval between prior infection and boosting of the immune response with a vaccine has been associated with more enhanced and durable immune responses.^16^ As shown in the present study, the evolution of post-vaccine immune responses is not even remotely close between those having experienced prior infection and the naïve. Non-stratified strategies for repeated boosters may lead to unexpected harms or attenuated performance through the ‘antibody feedback’ mechanism. Thus, when and whom to target with the repeated booster vaccinations remains a crucial question to all future vaccination campaigns. As long as rather young and/or healthy HCWs are targeted, it was shown in a preceding study from Israel that a third-dose vaccine was sufficient enough for totally preventing severe disease. The evidence here provided, that a fourth dose was only associated with a scaled-down additive protection of 39% reduction in infection risk, further prompts addressing this impending issue.^17^

The limitation of the study is the limited number of individuals evaluated. The observed immune response may not represent that of the overall population. The immune response of individuals from older age categories and at utmost risk of severe disease would have been highly intriguing, although not covered in the present study. The extreme elderly and multi-morbid population has been shown to exhibit aberrant immune responses.^18^

Hybrid immunity is becoming increasingly common. The benefits of boosting the infection-acquired immunity by vaccination has been shown ‘clinically’ to enhance the degree and duration of protection (protection rate persistently above 90% for 18 months or longer).^16^ The present study, in turn with robust indices of protective antibody response, further enriches the evidence for and provides an immunological basis to this highest and most durable protection achieved by those vaccinated on top of a primary infection. With hybrid immunity becoming increasingly prevalent, delayed boosters or reduced dosing regimens may become a realistic consideration when reshaping the future SARS-CoV-2 vaccination campaigns.

## Data Availability

All data produced in the present work are contained in the manuscript.

## Acknowledgments

The authors would like to thank all HCWs participating in the study. We would also like to thank Mrs. Mika Oku and Mrs. Takako Kobayashi from the Department of Virology & Parasitology, Graduate School of Medicine, Osaka Metropolitan University, for technical assistance in analyzing the specimens. Tomoyo Tominaga, Hiroko Tanaka, Tomoaki Yokoya, and Minako Hosokawa, from the Department of Health Management, St.Mariannna University, Yokohama Seibu Hospital, integrally supported the sample and data collection. Data acquisition with the microplate reader was performed at the Research Support Platform, Graduate School of Medicine, Osaka Metropolitan University.

